# Resident physician exposure to novel coronavirus (2019-nCoV, SARS-CoV-2) within New York City during exponential phase of COVID-19 pandemic: Report of the New York City Residency Program Directors COVID-19 Research Group

**DOI:** 10.1101/2020.04.23.20074310

**Authors:** Mark P. Breazzano, Junchao Shen, Aliaa H. Abdelhakim, Lora R. Dagi Glass, Jason D. Horowitz, Sharon X Xie, C. Gustavo de Moraes, Alice Chen-Plotkin, Royce W. S. Chen, on behalf of the New York City Residency Program Directors COVID-19 Research Group

## Abstract

**Background:** From March 2-April 12, 2020, New York City (NYC) experienced exponential growth of the COVID-19 pandemic due to novel coronavirus (SARS-CoV-2). Little is known regarding how physicians have been affected. We aimed to characterize COVID-19 impact on NYC resident physicians.

**Methods:** IRB-exempt and expedited cross-sectional analysis through survey to NYC residency program directors (PDs) April 3–12, 2020, encompassing events from March 2–April 12, 2020.

**Findings:** From an estimated 340 residency programs around NYC, recruitment yielded 91 responses, representing 24 specialties and 2,306 residents. 45.1% of programs reported at least one resident with confirmed COVID-19: 101 resident physicians were confirmed COVID-19-positive, with additional 163 residents presumed positive for COVID-19 based on symptoms but awaiting or unable to obtain testing. 56.5% of programs had a resident waiting for, or unable to obtain, COVID-19 testing. Two COVID-19-positive residents were hospitalized, with one in intensive care. Among specialties with >100 residents represented, negative binomial regression indicated that infection risk differed by specialty (p=0.039). Although most programs (80%) reported quarantining a resident, with 16.8% of residents experiencing quarantine, 14.9% of COVID-19-positive residents were not quarantined. 90 programs, encompassing 99.2% of the resident physicians, reported reuse or extended mask use, and 43 programs, encompassing 60.4% of residents, felt that personal protective equipment (PPE) was suboptimal. 65 programs (74.7%) have redeployed residents elsewhere to support COVID-19 efforts.

**Interpretation:** Many resident physicians around NYC have been affected by COVID-19 through direct infection, quarantine, or redeployment. Lack of access to testing and concern regarding suboptimal PPE are common among residency programs. Infection risk may differ by specialty.

**Funding:** AHA, MPB, RWSC, CGM, LRDG, JDH are supported by NEI Core Grant P30EY019007 and an unrestricted grant from RPB. ACP and JS are supported by the Parker Family Chair. SXX is supported by the University of Pennsylvania.

## INTRODUCTION

For the first time in over a century, the United States (US) is part of a global pandemic known as COVID-19,^1^ with anticipated impact comparable to the Spanish flu of 1918. The causative novel coronavirus (2019-nCoV, SARS-CoV-2), first described in Wuhan, China,^2,3^ has spread worldwide, particularly in New York City (NYC), which is currently the US epicenter of cases and mortality.^4^ The first case was confirmed in NYC on March 1, 2020;^5^ six weeks later, hundreds of patients are dying from COVID-19 daily.^6^ Healthcare workers (HCW) are on the front lines of this pandemic.^2,7^ However, although at least 4,500 peer-reviewed articles have been published on this topic between January 1, 2020 and April 18, 2020, comparatively little is known about the toll of COVID-19 on healthcare workers directly occupied with patient care.

Notably, the first physician to sound the alarm about the novel coronavirus causing severe acute respiratory syndrome coronavirus 2 (SARS-CoV-2) was the Chinese ophthalmologist Li Wenliang, who died after infection by a pre-symptomatic patient.^8^ Anecdotally, HCW in NYC have experienced unique challenges in combatting the illness, including close contact with the sickest patients, exposure to high viral loads, redeployment to clinical duties outside of their ordinary responsibilities, and severe shortages in personal protective equipment (PPE).^7,9,10^ Among those at highest risk are resident physicians, who are commonly stationed in high-acuity settings and comprise a substantial part of the healthcare workforce in the United States.^11^ The activities of resident physicians are standardized among residency training programs throughout the US via accreditation with the Accreditation Council for Graduate Medical Education (ACGME), with each residency program supervised by an appointed program director.^12^ The structure of residency programs, with many resident physicians reporting to one program director responsible for their activities and well-being, makes the resident physician population practical for study through collection of data from residency program directors. However, to our knowledge, no primary peer-reviewed data has addressed implications of COVID-19 for resident physicians, whose situation has only been described in editorials.^13,14^ We also sought to explore whether specialty-specific risks existed for COVID-19 infection. By surveying residency program directors among all departments within NYC from April 3-12, 2020, we captured the immediate features and impact of COVID-19 among resident physicians during the exponential phase of the COVID-19 pandemic in NYC. As future or recurrent outbreaks are likely, such knowledge may help tailor future interventions to mitigate the burden of COVID-19 among HCWs.

## METHODS

### Recruitment of program directors

Recruitment of residency program directors throughout the greater NYC area was performed through circulation of electronic mail message sent by one investigator at Columbia University Irving Medical Center (R.W.S.C.), with responses received from April 3, 2020 through April 12, 2020. Identification of programs, respective program directors, and contact electronic mail addresses were retrieved from either previous correspondence or publicly available search tools with ACGME via hyperlink (https://apps.acgme.org/ads/Public/Programs/Search). The survey was first distributed to 12 ophthalmology residency program directors in the greater NYC area, who expanded distribution to 188 additional non-ophthalmology training programs within their own institutions. As a second method, 303 programs identified separately in the ACGME database by two authors (M.P.B., A.H.A.) were also contacted electronically. Ultimately, at least one contact attempt was made at every known residency training program in the greater NYC area (approximately 340 total), as our two approaches may have overlapped.

### Survey of resident physician experience

An anonymous survey (**Supplemental Content**) eliciting de-identified information was included in circulated electronic mail message by hyperlink with SurveyMonkey® cloud-based software (SurveyMonkey®, San Mateo, CA, USA). More than one survey completion by the same user was prohibited, both by request within the recruitment electronic message and based on internet protocol address. The need for subject consent was waived due to minimal risk, anonymous nature, and lack of sensitive information in the study design as per Columbia University Institutional Review Board expedited exemption protocol IRB-AAAS9946. All procedures were reviewed and in accordance with the tenets of the Declaration of Helsinki.

Diagnosis or suspicion of COVID-19 among residents was elicited in our survey based on clinical presentation with symptoms including: sore throat, cough, fever, shortness of breath, chest pain, myalgia, malaise, conjunctivitis, anosmia, or gastrointestinal symptoms. Survey questions pertained to 3 distinct groups among resident doctors: (1) “confirmed” – defined as resident physicians with COVID-19 symptoms and positive test results; (2) “presumed” – defined as resident physicians with COVID-19 symptoms without test results, and (3) “suspected” – defined as resident physicians with COVID-19 symptoms and negative test results. Suspected cases were tallied in our analysis due to the relatively high false negative rate of reverse transcription polymerase chain reaction (RT-PCR) testing for active infection by this virus^15,16^ as well as high pre-test probability for COVID-19 in the context of suggestive symptoms, due to HCW status and NYC location.

### Inclusion and exclusion of responses

Responses were reviewed for inclusion based on specific training program. Fellowship programs were excluded from the analysis. Because certain specialties have programs that exist as a residency-fellowship continuum, these training programs with ACGME accreditation were included. We did not distinguish between these integrated programs and residency-only programs. All programs included were ACGME-accredited, with the exception of oral maxillofacial surgery (OMFS), which was included as many OMFS programs offer clinical experience through ACGME-accredited rotations such as general surgery, ultimately leading to medical licensure with or without an M.D. degree, in addition to pre-existing dental licensure. Programs were included if within or immediately adjacent to NYC. All queried programs but one were centralized within 30 miles of Central Park in Manhattan, verified by Google Maps with hyperlink: https://www.google.com/maps (Google Inc., Mountain View, CA, USA) for distance calculations which used mailing addresses from primary affiliations for each recipient of the survey.

### Statistical analyses

Proportions are reported as percentages with 95% confidence interval (CI) calculated using the Clopper-Pearson approach.

Specialties with representation by 100 or more residents were selected for further between-specialty analyses. Because the number of COVID-19 positive residents by individual programs were count outcomes and non-normally distributed, Poisson regression and negative binomial regression were fitted to determine whether specialty, program size, or date of survey response affected the number of residents with positive COVID-19 tests. Likelihood ratio (LR) testing was used to determine the appropriateness between Poisson regression and negative binomial regression. Fisher’s exact test was used to assess the overall effect of specialties on the proportion of residents with confirmed COVID-19. Pearson’s chi-squared test was used to compare infection rate and redeployment rate between departments. Correction for multiple comparisons was made with Bonferroni procedures.

Statistical analyses were performed in the R programming language (Version.1.2.5042). Type 1 error was defined at the 5% level for hypothesis testing with two-tailed probabilities.

## RESULTS

### Study sample

102 program director responses were received between April 3 and April 12, 2020, 10 of which were excluded based on residency and ACGME-accreditation criteria, and one of which was removed as it was incomplete and reported zero residents in the program. Thus, 91 programs representing 2,306 residents from 24 different specialties were included in this study (**Figure 1)**. Average program size was 25 residents (standard deviation [S.D.] = 21), with a range of 1 – 98 residents per program. 49 programs (53.8%, 95% CI 43.1-64.4) reported that residents provided services for ≤3 different hospitals.

**Figure 1.**
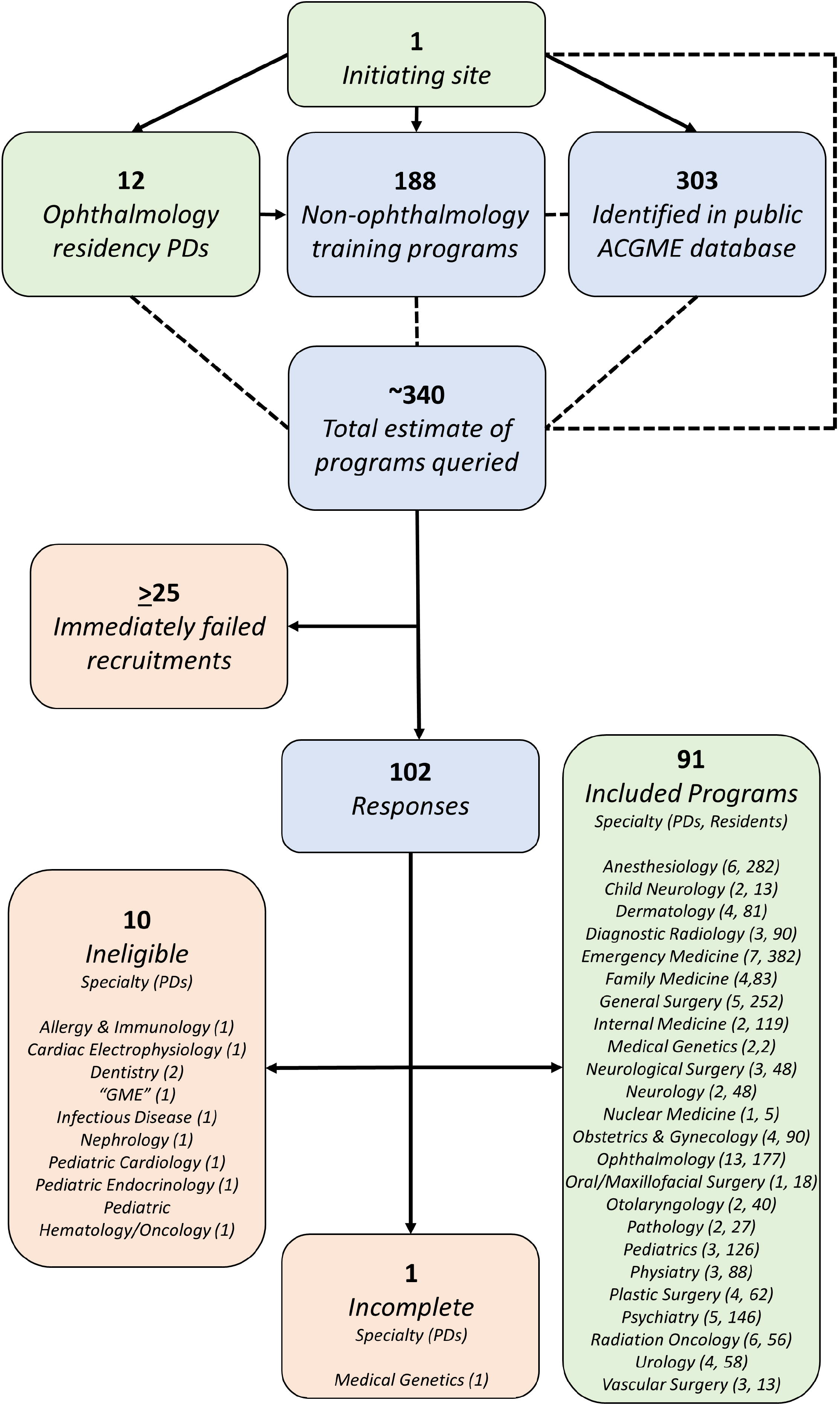
Flow-chart of survey recruitment and responses among greater New York City training programs, including represented specialties and number of residents. *ACGME = Accreditation Council for Graduate Medical Education; PDs = training program directors*.

### Overall cases and testing frequency of COVID-19

All 91 program directors reported numbers for symptomatic residents who had tested positive for COVID-19 (“confirmed” cases). 90/91 program directors reported numbers for symptomatic residents who were awaiting or unable to obtain testing (“presumed” cases) and symptomatic residents who had tested negative for COVID-19 (“suspected” cases). In total, 41/91 (45.1%, 95% CI 34.6-55.8) programs reported at least one confirmed case, 49/90 programs (54.4%, 95% CI 43.6-65.0) reported at least one presumed case, and 36/90 programs (40%, 95% CI 29.8-50.9) reported at least one suspected case. Among all residents from all programs pooled together, 101 residents were confirmed cases, 163 were presumed cases, and 76 were suspected cases (**Figure 2)**. The total number and proportion of affected residents by specialty are shown in the **Table**.

**Figure 2.**
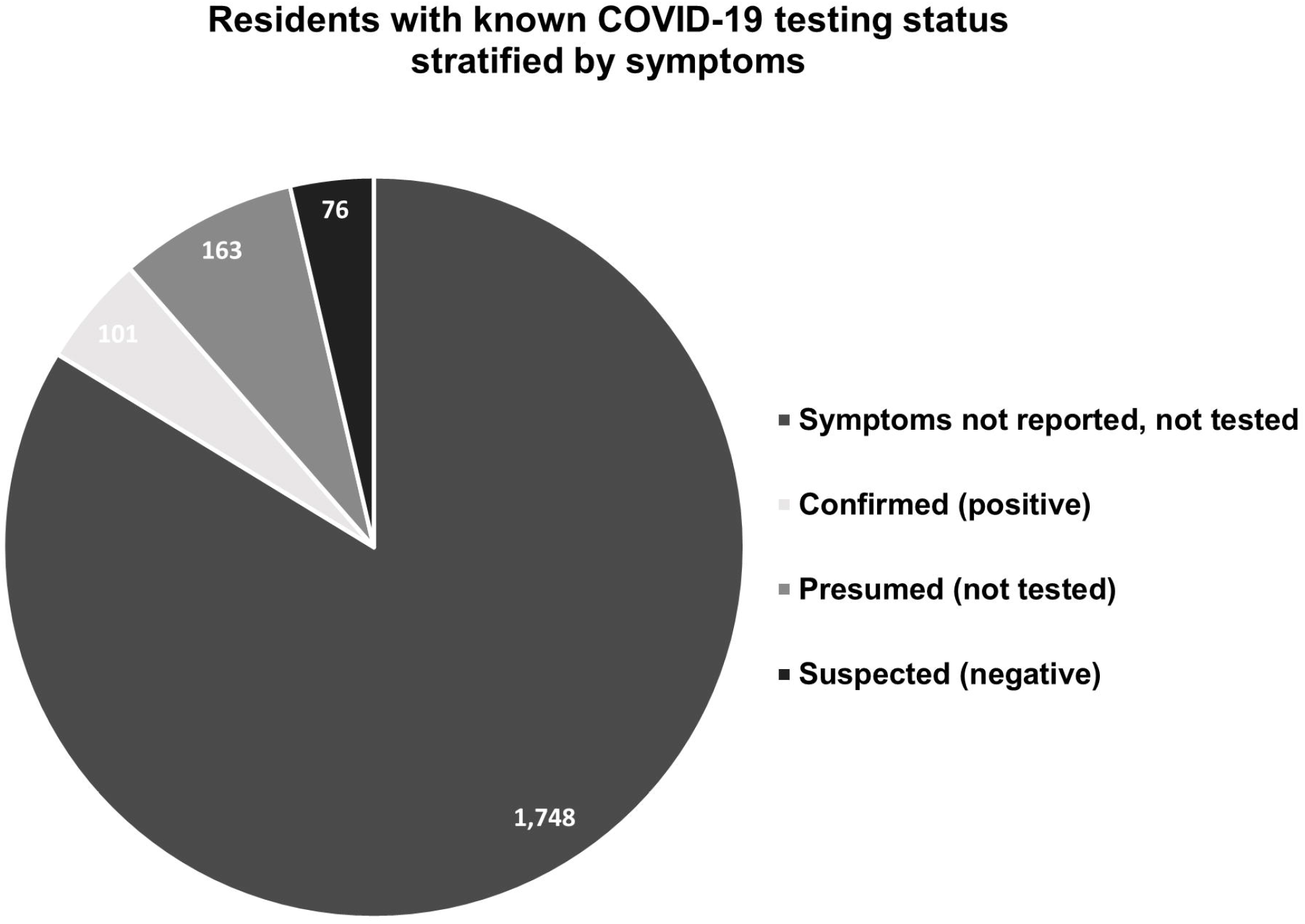
Of 2,088 total residents with known COVID-19 testing status, 101 residents were confirmed *(positive)*, 163 were presumed *(untested)*, 76 were suspected *(negative)*, and 1,748 neither had symptoms nor were tested.

86/91 program directors reported knowing how many residents were tested for COVID-19. Among the 2,088 residents in these 86 programs, a total of 242 residents (11.6%, 95% CI 10.2-13.0) were tested for COVID-19. 177 residents who were tested also had results reported by the time of the survey. Among these, 101 (57.1%, 95% CI 49.4%-64.5) tested positive and 76 (42.9%, 95% CI 35.5-50.6) were negative.

69/91 program directors reported knowing the exact number of residents who were tested for COVID-19 as well as whether residents were awaiting testing. Among 1,673 residents in these 69 programs, 113 residents (6.8%, 95% CI 5.6-8.1) were waiting for or unable to obtain testing. 39 (56.5%, 95% CI: 44.0-68.4) residency programs had at least 1 resident waiting for or unable to get testing.

For residents who tested positive for COVID-19 as well as those who tested negative, the majority of testing was performed with RT-PCR of samples collected by nasal swab (n=85 [84.2%] for test-positive; n=59 [77.6%] for test-negative), followed by oropharyngeal swab (n=5 [5.2%] for test-positive; n=6 [7.9%] for test-negative).

### Disease burden by specialty

To determine whether any specific medical specialties were more likely to have a COVID-19 positive resident, all specialties with more than 100 residents in our sample were compared. Programs that met this criterion included anesthesiology, emergency medicine, general surgery, internal medicine, ophthalmology, pediatrics, and psychiatry (**Figure 1**). Three specialties (anesthesiology, emergency medicine, ophthalmology) appeared to cluster as high-risk specialties by proportion of residents with confirmed COVID-19, compared to the remaining specialties (p=0.015, Fisher’s exact test). In negative binomial models adjusted for the size of the residency program and date of survey completion, specialty remained significantly associated with the number of confirmed positive residents (p= 0.039). Using anesthesiology as the reference group (as this specialty had the highest number of positive residents), anesthesiology was significantly more likely to have a COVID-19 confirmed resident, compared to both internal medicine (p= 0.020) and pediatrics (p = 0.029, **Figure 3**).

**Figure 3.**
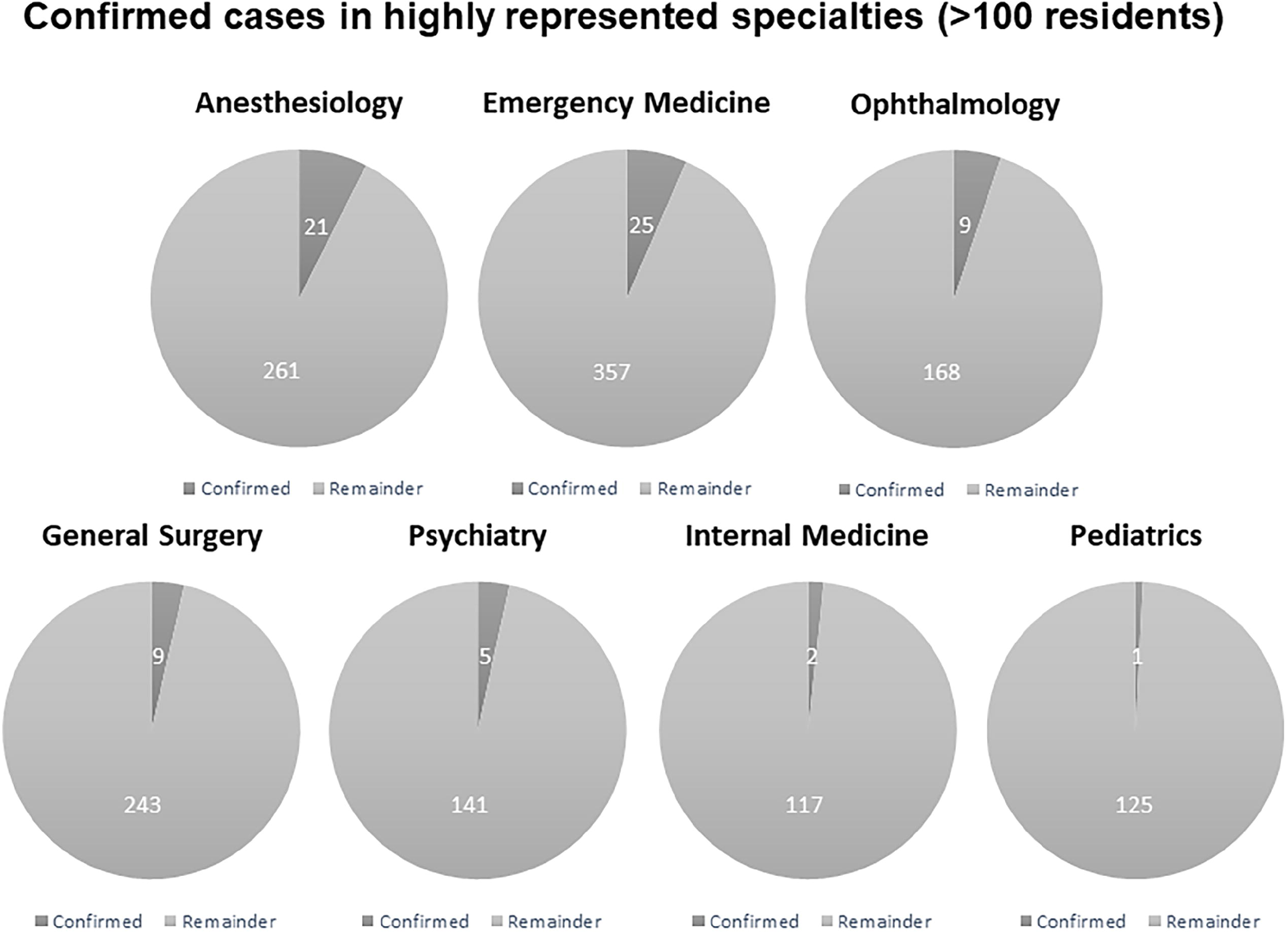
Number of residents by specialty with confirmed positive COVID-19 testing. All 7 specialties with a sample size greater than 100 residents as surveyed across 91 representative program directors among 24 specialties and 2,306 residents from the greater New York City area are included. Anesthesiology was significantly more likely to have a COVID-19 confirmed resident, compared to both internal medicine (p= 0.020) and pediatrics (p = 0.029).

### Timing of symptom onset

Symptom onset was reported to occur as early as or prior to the week of March 2–8, 2020 for 5 residents (1.5%) with confirmed (n=1), presumed (n=3), or suspected (n=1) COVID-19 (**Figure 4**). Most residents with confirmed COVID-19 (35, 34.7%, 95% CI 25.5-44.8) were reported to first experience symptoms the week of March 22–28, 2020. By contrast, most with presumed (53, 32.5%, 95% CI 25.4-40.3) and suspected (29, 38.2%, 95% CI 27.2-50.0) COVID-19 reported symptoms beginning the week of March 15–21, 2020. Symptom onset for affected residents among every category (confirmed: n=3 [3.0%], presumed: n=3 [1.8%], suspected: n=1 [1.3%]) continued through the last week of survey participation, April 6–12, 2020.

**Figure 4.**
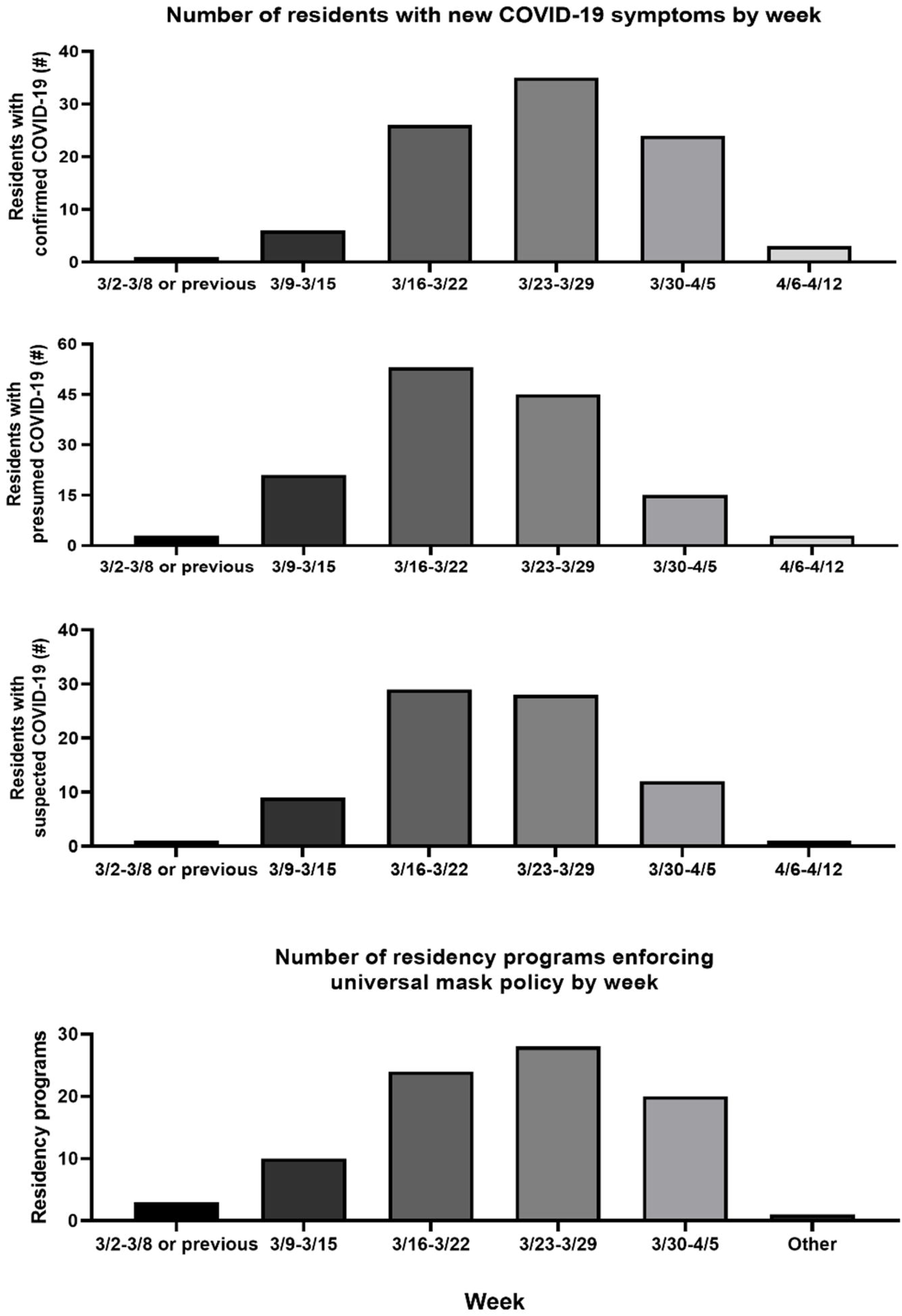
Number of residents with new COVID-19 symptoms by week. Most confirmed COVID-19 cases (N = 35) were reported during the week of 3/23-3/29. Most presumed COVID-19 cases (N= 53) and suspected COVID-19 cases (N = 29) were reported a week earlier than the peak of confirmed cases during 3/16 – 3/22. Total number of confirmed, presumed and suspected COVID-19 cases all started to drop after the week of 3/23 -3/29. Bottom panel shows the number of programs enforcing mask policy by week. Most programs started to enforce universal mask policy during the week of 3/23 – 3/29.

### Personal protective equipment (PPE)

The majority of programs, encompassing 1,832 residents (79.4%, 95% CI 77.7-81.1) used either N95 or surgical masks during patient encounters, depending on the context. Nineteen programs, encompassing 323 residents (14%, 95% CI 12.6-15.5) used only surgical masks during patient encounters; and 8 programs, encompassing 31 residents, (5.7%, 95% CI 4.8-6.7) used an N95 respirator for all patient encounters. Excepting one radiology program, all programs, encompassing 99.2% of residents in this study, reported reuse or extended use of their masks (vs. single-use). Protocols mandating universal wearing of surgical masks were introduced as early as the week of March 2–8, 2020 in only 3 programs (3.5%), and as late as March 30–April 5, 2020 in 20 programs (23.5%, **Figure 4**).

43/87 program directors (49.4%, 95% CI 38.5-60.4) representing 1,314 residents answered “yes” when asked whether their residents had had to work with suboptimal PPE. There was no correlation between the mask type used by residents (surgical, N95, or both) to perceived shortage of PPE.

### Care Setting and Hospitalization

Among the 101 residents with confirmed COVID-19, 57 (56.4%, 95% CI 46.2-66.3) presented to clinic or primary care, 17 (16.8%, 95% CI 10.1-25.6) visited the emergency department, 2 (2.0%, 95% CI 0.2-7.0) were hospitalized, and 1 (1%, 95% CI 0-5.4) had care escalated to the intensive care unit (ICU). The 163 residents with presumed COVID-19 presented to primary care or clinic in 40 cases (24.5%, 95% CI 18.1-31.9) and the emergency department in 6 cases (3.7%, 95% CI 1.4-7.8). Among the 76 residents with suspected COVID-19, 38 (50%, 95% CI 38.3-61.7) were evaluated in clinic or by primary care, 5 (6.5%, 95% CI 2.2-14.7) presented to emergency department, and 1 (1.3%, 95% CI 0-7.1) was hospitalized. In total, among the 340 residents with confirmed, presumed or suspected COVID-19, 3 (0.9%, 95% CI 0.2-2.6) were hospitalized (1 each from emergency medicine [who was also hospitalized and went to the ICU], ophthalmology, and psychiatry programs; 2 were confirmed, and 1 suspected COVID-19). There were no deaths reported in any of the completed surveys.

### Quarantine

One program (pediatrics) of 58 residents did not report any quarantine data. Of the remaining 90 programs encompassing 2,248 residents (including 339 residents with confirmed, presumed, or suspected COVID-19 infection), 377 (16.8%, 95% CI 15.2-18.4) residents from 72 programs (80% of programs, 95% CI 70.2-87.7) were reported to be quarantined. 22 programs (24.4%, 95% CI 16.0-34.6) reported at least one asymptomatic, but exposed, resident, who was quarantined. Among 34 asymptomatic but exposed residents with known duration of quarantine, the time ranged from 1 – 14 days. 15 residents (14.9%, 95% CI 8.6-23.3) from 2 programs with confirmed COVID-19, 26 residents (16.0%, 95% CI 10.8-22.6) from 5 programs with presumed COVID-19, and 5 residents (6.6%, 95% CI 2.2-14.7) from 2 programs with suspected COVID-19 were not quarantined.

### Redeployment

87/91 program directors responded to questions about residents redeployed to other departments or locations to support COVID-19 efforts. 65 programs (74.7%, 95% CI 64.3-83.4) reported at least one resident redeployed, with 35 (40.2%, 95% CI 29.9-51.3) programs redeploying more than one-third of their workforce. 594 residents (27.3% of 2,176 residents for whom redeployment information is known, 95% CI 25.4-29.2) were reported to be redeployed. Anesthesiology had the highest redeployment rate, with 158 (56.0% of 282 total anesthesiology residents, 95% CI 50.0-61.9) residents being redeployed to other services (*p*<0.001, Pearson’s chi-squared test). Of programs that redeployed residents, 53 programs (81.5%, 95% CI 70.0-90.1) instituted redeployment between the fourth and fifth weeks of March, approximately 1 month after the first case in NYC was confirmed. Among residents redeployed to duties beyond their usual clinical responsibilities, the majority went to the ICU (283/594 redeployed residents, 47.6%, 95% CI 43.6-51.7), followed by hospital floors (176/594, 29.6%, 95% CI 26.0-33.5), and the emergency department (85/594, 14.3%, 95% CI 11.6-17.4).

## DISCUSSION

As of the date of our survey’s close, NYC is the epicenter of the COVID-19 pandemic in the US, and the daily death toll continues to rise.^6^ Here, we report the impact of COVID-19 on NYC resident physicians, as reported by their residency program directors, surveyed between April 3-12, 2020. Many of these residents have been directly infected (101 confirmed positive), quarantined (16.8% of residents), or redeployed (27.3% of residents) to duties outside of their usual clinical activities in support of COVID-19 efforts.

101 residents were reported to have confirmed COVID-19 in our sample. While this is 4.4% of the 2306 residents whose program directors participated in our study, the true rate in our sample may be higher, since 242 resident physicians were tested for COVID-19, and only 177 had received their test results at the close of the survey.

We highlight a few points found in our study. First, program directors reported 15 confirmed COVID-19 residents and 26 presumed COVID-19 residents who were not quarantined. Whether this was due to these residents being initially asymptomatic, workforce need, delay in obtaining testing, or some other reason is not known.

However, we do note that 57.4% of residency program directors reported at least one resident awaiting or unable to obtain COVID-19 testing. Second, 49.4% of residency directors answered “yes” to the question of whether resident physicians for whom they were responsible had suboptimal PPE. While this might reflect selection bias with respect to which residency directors chose to answer the survey, we note that 90/91 programs reported reuse or extended use of masks that are ordinarily disposable after a single use. Third, we find that some specialties may be at greater risk for contracting COVID-19 compared to others. In particular, anesthesiology had significantly higher numbers of confirmed COVID-19 residents than several other specialties. It is possible that the higher infection rates may be due to the critical skill of intubation provided by anesthesiologists, which comes with high probability of aerosolization and exposure to viral particles.^17^

We recognize limitations to our current study. While not all presumed and suspected cases have COVID-19, we present these numbers given the high pre-test probability of infection in HCW with suggestive symptoms, as well as known limitations of RT-PCR detection of the virus.^15,16^ Future work using serological testing may provide a more accurate census of confirmed positives, as recent studies have shown.^18^ Selection bias may have affected our findings, as fields such as ophthalmology may have been over-represented due to the authors’ connections to colleagues in this field, while other specialties may have been under-represented because of significant stress and lack of time to complete the survey. It is also possible that program directors whose residents have been affected by COVID-19 would be more likely to respond. However, we capture 91 NYC residency programs (out of an estimated 340 total residency programs) during a period of exponential pandemic growth, offering a unique perspective on the impact on resident physicians during what may be the height of COVID-19 in NYC. Indeed, capturing the experience as it happens avoids recall bias after the fact. It is our hope that this insight may allow locations not yet as substantially affected by COVID-19 to better anticipate the needs of resident physicians, who are truly at the front lines of an unprecedented challenge.

### New York City Residency Program Directors COVID-19 Research Group

Azin Abazari, MD^6^, Douglas R. Frederick, MD^7^, Ilana B. Friedman, MD^8^, Albert S. Khouri, MD^9^, Eleanore T. Kim, MD^2^, John J. Laudi, MD^10^, Jamie B. Rosenberg, MD^11^, Harsha S. Reddy, MD^12^, Grace Sun, MD^13^, Jules Winokur, MD^3^, A. Elisabeth Abramowicz, MD, FASA^14^, Todd A. Anderson, MD^15^, Melissa Arbuckle, MD, PhD^16^, Judith Aronsohn, MD^17^, Teresa Benacquista, MD^18^, John Bent, MD^19^, Russell S. Berman, MD^20^, Jeremy Branzetti, MD^21^, Jeffrey N. Bruce, MD, FACS^22^, Eric D. Brumberger, MD^23^, Judah Burns, MD^24^, Simon Cheng, MD, PhD^25^, Michael A. Curi, MD, MPA^26^, Kevin Du, MD, PhD, MSCI^27^, Mary Fatehi, MD^28^, Mirela Feurdean, MD, FACP^29^, Michael A. Feuerstein, MD^30^, Sharon A. Glick, MD, FAAD, FAAP^31^, Karen E. Gibbs, MD^32^, Ira M. Goldstein, MD, FAANS^33^, Heidi J. Hansen, DMD^34^, Basil Hubbi, MD^35^, Machteld E. Hillen, MD^36^, Alejandro D. Iglesias, MD^37^, Michael P. Jones, MD^38^, Ana H. Kim, MD^39^, Anastasia Kunac, MD^40^, Renée M. Moadel, MD^24^, Peter Muscarella II, MD^41^, Aaron Nelson, MD, MBS, FAAP^42^, Heather L. Paladine, MD^43^, Yar Pye, MD, MBA^44^, Rini Banerjee Ratan, MD^45^, Matthew S. Robbins, MD, FAAN, FAHS^46^, Marsha E. Rubin, DDS, MAGD, FAOM, DABSCD^34^, Nicholas J. Sanfilippo, MD^47^, Helen L. Schleimer, MD^16^, Michael Schulder, MD, FAANS^48^, Andrew D. Schweitzer, MD^49^, Devorah Segal, MD, PhD^50^, Harsimran S. Singh, MD, MSc^51^, Harsh Sule, MD, MPP^52^, Peter J. Taub, MD, MS^53^, Lizica Troneci, MD^54^, Jonathan P. Ungar, MD, FAAD^55^, Patricia M. Vuguin, MD^37^, Michael Wajda, MD^56^, David Weithorn, MD^42^, Alan H. Wolff, MD^29^, Saqib Zia, MD, FACS^26^.

Affiliations: ^6^Department of Ophthalmology, State University of New York at Stony Brook Health Sciences Center, Stony Brook, NY, USA; ^7^Department of Ophthalmology, Icahn School of Medicine at Mount Sinai, New York, NY, USA; ^8^Department of Ophthalmology, BronxCare Health System, Bronx, NY, USA; ^9^Department of Ophthalmology & Visual Science, Rutgers – New Jersey Medical School, University Hospital, Newark, NJ, USA; ^10^Department of Ophthalmology, State University of New York Downstate Health Sciences University, Brooklyn, NY, USA; ^11^Department of Ophthalmology & Visual Sciences, Albert Einstein College of Medicine, Montefiore Medical Center, New York, NY, USA; ^12^Department of Ophthalmology, New York Eye and Ear Infirmary of Mount Sinai, New York, NY, USA; ^13^Department of Ophthalmology, Weill Medical College of Cornell University, New York-Presbyterian Hospital, New York, NY, USA; ^14^Department of Anesthesiology, Westchester Medical Center, New York Medical College, Valhalla, NY, USA; ^15^Lenox Hill Pathology, Zucker School of Medicine at Hofstra, Northwell Health, New York, NY, USA; ^16^New York State Psychiatric Institute, Department of Psychiatry, Columbia University Irving Medical Center, New York-Presbyterian Hospital, New York, NY, USA; ^17^Department of Anesthesiology, Zucker School of Medicine at Hofstra, Northwell Health, New York, NY, USA; ^18^Department of Plastic & Reconstructive Surgery, Albert Einstein College of Medicine, Montefiore Medical Center, Bronx, NY, USA; ^19^Department of Otolaryngology, Albert Einstein College of Medicine, The Children’s Hospital at Montefiore, Bronx, NY, USA; ^20^Department of Surgery, New York University School of Medicine, New York University Langone Health, New York, NY, USA; ^21^Department of Emergency Medicine, New York University School of Medicine, New York University Langone Health, New York, NY, USA; ^22^Department of Neurological Surgery, Bartoli Tumor Research Laboratory, Columbia University Irving Medical Center, New York-Presbyterian Hospital, New York, NY, USA; ^23^Department of Anesthesiology, Weill Cornell Medical College, New York-Presbyterian Hospital, New York, NY, USA; ^24^Department of Radiology, Albert Einstein College of Medicine, Montefiore Medical Center, New York, NY, USA; ^25^Department of Radiation Oncology, Columbia University Irving Medical Center, New York-Presbyterian Hospital, New York, NY, USA; ^26^Department of Vascular Surgery, Rutgers – New Jersey Medical School, University Hospital, Newark, NJ, USA; ^27^Department of Radiation Oncology, New York University School of Medicine, New York University Langone Health, New York, NY, USA; ^28^Department of Obstetrics & Gynecology, Nassau University Medical Center, New York, NY, USA; ^29^Department of Medicine, Rutgers – New Jersey Medical School, University Hospital, Newark, NJ, USA; ^30^Department of Urology, Lenox Hill Hospital, Zucker School of Medicine at Hofstra, Northwell Health, New York, NY, USA; ^31^Department of Dermatology, State University of New York Downstate Health Sciences, Brooklyn, NY, USA; ^32^Department of Surgery, Staten Island University Hospital, Zucker School of Medicine at Hofstra, Northwell Health, New York, NY, USA; ^33^Department of Neurological Surgery, Rutgers – New Jersey Medical School, Newark Beth Israel Medical Center, Newark, NJ, USA; ^34^Department of Surgery, Division of Dentistry, Oral and Maxillofacial Surgery, Weill Cornell Medical College, New York-Presbyterian Hospital, New York, NY, USA; ^35^Department of Radiology, Rutgers – New Jersey Medical School, University Hospital, Newark, NJ, USA; ^36^Department of Neurology, Rutgers – New Jersey Medical School, University Hospital, Newark, NJ, USA; ^37^Department of Pediatrics, Columbia University Irving Medical Center, New York-Presbyterian Hospital, New York, NY, USA; ^38^Department of Emergency Medicine, Albert Einstein College of Medicine, Montefiore Medical Center, New York, NY, USA; ^39^Department of Otolaryngology – Head & Neck Surgery, Columbia University Irving Medical Center, New York-Presbyterian Hospital, New York, NY, USA; ^40^Department of Surgery, Rutgers – New Jersey Medical School, University Hospital, Newark, NJ, USA; ^41^Department of Surgery, Albert Einstein College of Medicine, Montefiore Medical Center, New York, NY, USA; ^42^Department of Neurology, New York University School of Medicine, New York University Langone Health, New York, NY, USA; ^43^Center for Family and Community Medicine, Columbia University Irving Medical Center, New York-Presbyterian Hospital, New York, NY, USA; ^44^Department of Medicine, New York University School of Medicine, New York University Langone Health, New York, NY, USA; ^45^Department of Obstetrics & Gynecology, Columbia University Irving Medical Center, New York-Presbyterian Hospital, New York, NY, USA; ^46^Department of Neurology, Weill Cornell Medical College, New York-Presbyterian Hospital, New York, NY, USA; ^47^Department of Radiation Oncology, Weill Cornell Medical College, New York-Presbyterian Hospital, New York, NY, USA; ^48^Department of Neurological Surgery, Neurosciences Brain Tumor Center, Zucker School of Medicine, Northwell Health, New York, NY, USA; ^49^Department of Radiology, Weill Cornell Medical College, New York-Presbyterian Hospital, New York, NY, USA; ^50^Department of Pediatrics, Weill Cornell Medical College, New York-Presbyterian Hospital, New York, NY, USA; ^51^Department of Anesthesiology, Weill Cornell Medical College, New York-Presbyterian Hospital, New York, NY, USA; ^52^Department of Emergency Medicine, Rutgers – New Jersey Medical School, University Hospital, Newark, NJ, USA; ^53^Department of Surgery, Icahn School of Medicine at Mount Sinai, New York, NY, USA; ^54^Department of Psychiatry, St. Barnabas Hospital Health System, Bronx, NY, USA; ^55^Department of Dermatology, Icahn School of Medicine at Mount Sinai, New York, NY, USA; ^56^Department of Anesthesiology, New York University School of Medicine, New York University Langone Health, New York, NY, USA

## Data Availability

Data are available upon request when and if appropriate.

## ACKNOWLEDGMENTS

We thank Julia A. Kucherich, RD, for contributing to survey design and discussion references.

---

**Table.** Number and percentage of symptomatic residents with confirmed *(positive)*, presumed *(untested)*, and suspected *(negative)* COVID-19 testing across specialties.

|  | # Residents | # Confirmed | # Presumed | # Suspected |
| --- | --- | --- | --- | --- |
| Vascular Surgery | 13 | 4 (30.8%) | 1 (7.7%) | 0 (0.0%) |
| Anesthesiology | 282 | 21 (7.4%) | 19 (6.7%) | 12 (4.3%) |
| Emergency Medicine | 382 | 25 (6.5%) | 32 (8.4%) | 12 (3.1%) |
| Radiation Oncology | 56 | 3 (5.4%) | 3 (5.4%) | 2 (3.6%) |
| Ophthalmology | 177 | 9 (5.1%) | 17 (9.6%) | 7 (4.0%) |
| Otolaryngology | 40 | 2 (5.0%) | 3 (7.5%) | 2 (5.0%) |
| Plastic Surgery | 62 | 3 (4.8%) | 17 (27.4%) | 0 (0.0%) |
| Physical Medicine and Rehabilitation | 88 | 4 (4.5%) | 4 (4.5%) | 1 (1.1%) |
| Obstetrics and Gynecology | 90 | 4 (4.4%) | 7 (7.7%) | 1 (1.1%) |
| Dermatology | 81 | 3 (3.7%) | 1 (1.2%) | 7 (9.2%) |
| Pathology | 27 | 1 (3.7%) | 1 (3.7%) | 0 (0.0%) |
| General Surgery | 252 | 9 (3.6%) | 16 (6.3%) | 13 (5.2%) |
| Psychiatry | 146 | 5 (3.4%) | 10 (6.8%) | 5 (3.4%) |
| Family Medicine | 83 | 2 (2.4%) | 3 (3.6%) | 7 (8.4%) |
| Neurological Surgery | 48 | 1 (2.1%) | 1 (2.1%) | 4 (8.3%) |
| Neurology | 48 | 1 (2.1%) | 4 (8.3%) | 1 (2.1%) |
| Internal Medicine | 119 | 2 (1.7%) | 5 (4.2%) | 0 (0.0%) |
| Diagnostic Radiology | 90 | 1 (1.1%) | 5 (5.6%) | 0 (0.0%) |
| Pediatrics | 126 | 1 (0.7%) | 2 (1.6%) | 0 (0.0%) |
| Urology | 58 | 0 (0.0%) | 7 (12.1%) | 0 (0.0%) |
| Child Neurology | 13 | 0 (0.0%) | 4 (30.8%) | 1 (8.6%) |
| Nuclear Medicine | 5 | 0 (0.0%) | 1 (20.0%) | 1 (20.0%) |
| Oral/Maxillofacial Surgery | 18 | 0 (0.0%) | 0 (0.0%) | 0 (0.0%) |
| Medical Genetics | 2 | 0 (0.0%) | 0 (0.0%) | 0 (0.0%) |
| **Total** | **2,306** | **101 (4.4%)** | **163 (7.1%)** | **76 (3.3%)** |

